# Three doses of COVID-19 mRNA vaccination are safe based on adverse events reported in electronic health records

**DOI:** 10.1101/2021.11.05.21265961

**Authors:** Michiel J.M. Niesen, Colin Pawlowski, John C. O’Horo, Doug W. Challener, Eli Silvert, Greg Donadio, Patrick J. Lenehan, Abinash Virk, Melanie D. Swift, Leigh L. Speicher, Joel Gordon, Holly L. Geyer, John Halamka, AJ Venkatakrishnan, Venky Soundararajan, Andrew Badley

## Abstract

Recent reports on waning of COVID-19 vaccine induced immunity have led to the approval and roll-out of additional dose and booster vaccinations. At risk individuals are receiving additional vaccine dose(s), in addition to the regimen that was tested in clinical trials. The risks and the adverse event profiles associated with these additional vaccine doses are currently not well understood. Here, we performed a retrospective study analyzing vaccine-associated adverse events using electronic health records (EHRs) of individuals that have received three doses of mRNA-based COVID-19 vaccines (n = 47,999). By comparing symptoms reported in 2-week time periods after each vaccine dose and in a 2-week period before the 1^st^ vaccine dose, we assessed the risk associated with 3^rd^ dose vaccination, for both BNT162b2 and mRNA-1273. Reporting of severe adverse events remained low after the 3^rd^ vaccine dose, with rates of pericarditis (0.01%, 0%-0.02% 95% CI), anaphylaxis (0.00%, 0%-0.01% 95% CI), myocarditis (0.00%, 0%-0.01% 95% CI), and cerebral venous sinus thrombosis (no cases), consistent with earlier studies. Significantly more individuals (p-value < 0.05) report low-severity adverse events after their 3^rd^ dose compared with after their 2^nd^ dose, including fatigue (4.92% after 3^rd^ dose vs 3.47% after 2^nd^ dose), lymphadenopathy (2.89% vs 2.07%), nausea (2.62% vs 2.04%), headache (2.47% vs 2.07%), arthralgia (2.12% vs 1.70%), myalgia (1.99% vs 1.63%), diarrhea (1.70% vs 1.24%), fever (1.11% vs 0.81%), vomiting (1.10% vs 0.80%), and chills (0.47% vs 0.36%). Our results show that although 3^rd^ dose vaccination against SARS-CoV-2 infection led to increased reporting of low-severity adverse events, risk of severe adverse events remained comparable to the standard 2-dose regime. This study provides support for the safety of 3^rd^ vaccination doses of individuals that are at high-risk of severe COVID-19 and breakthrough infection.

## Introduction

Although clinical trials^1,2^ and early studies^3,4^ have shown high effectiveness of mRNA-based COVID-19 vaccines at preventing SARS-CoV-2 infection and reducing COVID-19 severity, several recent studies suggest that vaccine effectiveness against milder disease is waning.^5–9^ This likely owes to waning immunity, as well as poorer immune response in certain at risk groups such as the immune compromised. ^10,11^ This has prompted health policy discussions on the need for additional/booster vaccine doses.

Initial studies indeed show that 3^rd^ dose vaccination results in improved protection against SARS-CoV-2 infection.^12,13^ An additional vaccine dose might be particularly beneficial for individuals at high risk of breakthrough infections or severe COVID-19, such as immunocompromised patients, as suggested by measurement of antibody levels in adults with solid tumors.^14^ However, despite these potential benefits, it is essential to monitor the safety of additional vaccine doses beyond the primary series as they are administered to the general public.

Extensive research has shown that the standard 2-dose regimen of mRNA-based COVID-19 vaccines is relatively safe. Although severe adverse events such as anaphylaxis,^15^ myocarditis,^16,17^ and blood clotting,^18–22^ have been reported after COVID-19 vaccination, these are rare and the benefit of vaccination is deemed to outweigh the potential risks. The most common adverse reactions occur immediately post vaccination^23^ and are relatively mild, including headache, fatigue, pains, low-grade fever, and nausea. Initial studies, using clinical trials,^13,24^ self-reporting of adverse events in a small cohort of 3^rd^ vaccine dose recipients,^25^ and analyzing voluntary reports in v-safe,^26^ indicate additional vaccination doses might also be safe, although further study in a broader study population is needed.

In addition to the existing studies, real-world evidence extracted from electronic health records (EHR) can help confirm the safety of COVID-19 additional vaccination doses. The advantage of this method for collecting adverse event reports is that a large and diverse cohort of individuals can be readily included, due to avoiding barriers associated with study enrollment or self-reporting. However, successful extraction of vaccine-associated adverse events from EHR data is challenging due to the vast amount of data involved and the difficulty in verifying that mention of a symptom corresponds with the patient experiencing the symptom at a specific point in time.

In this study, we aim to determine the safety of additional vaccine doses for individuals that previously received the standard 2-dose regimen of a mRNA-based COVID-19 vaccine. We evaluate vaccine-associated symptoms reported in cohorts of individuals that have received additional vaccination with either BNT162b2 or mRNA-1273.

## Methods

### Study population

This is a retrospective study of individuals, within the Mayo Clinic Enterprise who were vaccinated with exactly three doses of FDA-approved mRNA-based COVID-19 vaccines between December 1, 2020, and October 17, 2021. This study was reviewed and determined exempt by the Mayo Clinic Institutional Review Board. Individuals who had specifically opted out of inclusion of electronic medical records in research were excluded. Inclusion and exclusion criteria were defined as follows:

1. Age greater than or equal to 18 years at the date of initial COVID-19 vaccination.
2. Received the first two doses of BNT162b2 or mRNA-1273 per-emergency use authorization protocol. Per-protocol BNT162b2 vaccination is defined as two BNT162b2 doses administered 18-28 days apart. Per-protocol mRNA-1273 vaccination is defined as two mRNA-1273 doses administered 25-35 days apart.
3. Received a 3^rd^ dose of an mRNA-based COVID-19 vaccine at least 28 days after the 2^nd^ dose.
4. 3^rd^ COVID-19 vaccine dose of the same type as the original 2 doses.
5. Did not have more than three doses of an mRNA-based COVID-19 vaccine on record.
6. Did not previously receive any doses of a non-mRNA based COVID-19 vaccine (e.g. Janssen - Ad26.COV2.S).
7. At least 14 days of follow up after their 3^rd^ vaccine dose.

Study participants are divided into four cohorts for analysis, depending on the vaccine types of their initial two vaccine doses and their 3^rd^ vaccine dose (**Figure 1a**). Specifically, cohorts of individuals with three BNT162b2 vaccine doses (n = 38,094) and three mRNA-1273 vaccine doses (n = 9,905). Demographic and clinical characteristics of the cohorts are shown in **Table S1**, and information on the timing of vaccine doses is shown in **Figure S1**.

**Figure 1:**
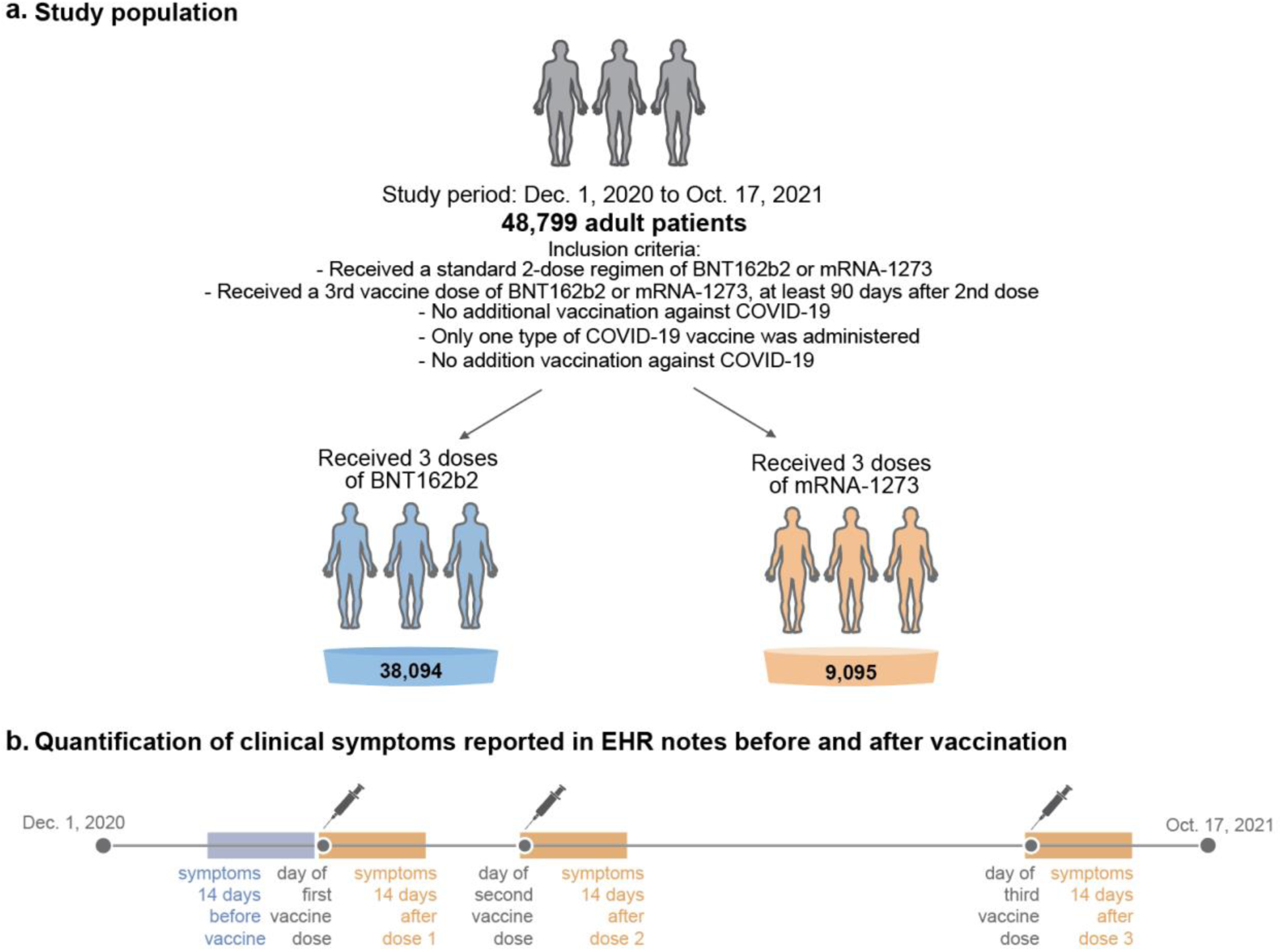
Schematic depiction of the study design. (**a**) The study population consists of 48,799 adult patients at Mayo Clinic Health System that meet all study inclusion criteria. This study population is subdivided into cohorts depending on the vaccine doses they received. (**b**) To find significant adverse events associated with COVID-19 vaccination, we quantified clinical symptoms reported in EHR notes during 14-day periods starting at the date of each vaccine dose (orange), as well as a control 14-day period before the 1^st^ vaccine dose (blue).

### Extracting adverse event sentiments from EHR data using augmented curation

A BERT-based^27^ classification model was used to determine the sentiment of adverse event phenotypes mentioned in the clinical notes. This model has been previously used to identify signs and symptoms of COVID-19,^28^ short and long-term complications of COVID-19,^29^ and adverse events of mRNA-based COVID-19 vaccines.^23^ Given a sentence which includes a phenotype, this model outputs one of the following labels: *Yes* - confirmed diagnosis, *Maybe* - possible diagnosis, *No* - ruled out diagnosis, or *Other* - none of the above (e.g. family history of diagnosis). This model was trained on a dataset of 18,490 sentences from clinical notes in the Mayo Clinic including over 250 different phenotypes and achieves an out-of-sample accuracy of 93.6% and precision and recall values above 95%.^28^

For this analysis, the above classification model was applied to clinical notes of each of the individuals in the study population for the following time periods: -15 to -1 days prior to the 1^st^ vaccine dose, 0 to 14 days following the 1^st^ vaccine dose, 0 to 14 days following the 2^nd^ vaccine dose, and 0 to 14 days following the 3^rd^ vaccine dose (**Figure 1b**). 19 adverse event phenotypes were considered, including: anaphylaxis, arthralgia, cerebral venous sinus thrombosis (CVST), chills, diarrhea, erythema, facial paralysis, fatigue, fever, headache, local pain, local swelling, lymphadenopathy, myalgia, myocarditis, nausea, pericarditis, soreness, and vomiting. For each (adverse event, time period) pair, individuals with at least one clinical note labelled “Yes” by the model with over 90% confidence were counted as having the adverse event. For a select set of rare severe adverse events (anaphylaxis, facial paralysis, myocarditis, pericarditis), additional manual curation was performed (by JCO and DWC) to confirm that the patients identified by the model did experience the adverse events during the time period of interest and that the adverse event was not attributed to another known cause (i.e., anaphylaxis due to allergic reaction to a known non-vaccine allergen).

### Estimation of adverse event risk from EHR data

We used previously described augmented curation models to extract sentiments of adverse events from clinical notes.^23,28^ Specifically, for each individual in a cohort, we determine whether positive sentiments for vaccine-associated adverse events are present in their clinical notes, during a specific 14-day period relative to their date of vaccination (**Figure 1**). The risk of an adverse event is then reported as the percentage of vaccine recipients in a cohort with a positive sentiment for that adverse event. This risk is compared to the baseline risk in the cohort, taken as the risk for the adverse event in a 14-day period before the 1^st^ COVID-19 vaccine dose. The reported confidence intervals and p-values are determined using bootstrap resampling (N=10,000 samples).

### Extracting comorbidity and immunosuppressant medication data from EHRs

For each patient, Elixhauser comorbidities^30^ for HIV/AIDS and cancer were determined using ICD-10 codes in the 5 year period leading up to the 1^st^ COVID-19 vaccine dose. In addition, overall Elixhauser comorbidity scores were computed using the Van Walraven method.^31^ Patients who had taken immunosuppressant medications in the past 1 year in their medical history were identified by querying the EHR database for the list of medications in the drug class WHO ATC LO4A^.32^

### IRB approval for human subjects research

This study was reviewed and approved by the Mayo Clinic Institutional Review Board (IRB 20 - 003278) as a minimal risk study. Subjects were excluded if they did not have a research authorization on file. The approved IRB was titled: Study of COVID-19 patient characteristics with augmented curation of Electronic Health Records (EHR) to inform strategic and operational decisions with the Mayo Clinic. The following resource provides further information on the Mayo Clinic Institutional Review Board and adherence to basic ethical principles underlying the conduct of research, and ensuring that the rights and well-being of potential research subjects are adequately protected: www.mayo.edu/research/institutional-review-board/overview.

## Results

Overall, we find no significant difference in the reporting of severe adverse events and a significant increase in reporting for most low-severity adverse events after the 3^rd^ COVID-19 vaccine dose, compared with earlier doses.

The most common adverse events reported after the 3^rd^ vaccine dose were fatigue (4.92%), lymphadenopathy (2.89%), nausea (2.62%), headache (2.47%), arthralgia (2.12%), myalgia (1.99%), diarrhea (1.70%), erythema (1.00%), fever (1.11%), vomiting (1.10%), chills (0.47%), and soreness (0.36%) (**Figure S2**). The median time between the vaccine dose and reporting of the adverse events is listed in **Table S2** and **Table S3**. We quantified increased reporting of adverse events after the 3^rd^ dose, compared with the 2^nd^ dose and baseline incidence, using the risk difference (RD). We found that compared to the 2^nd^ dose, there was increased reporting of most common adverse events for both BNT162b2 and mRNA-1273 after dose #3. Notably, overall patients reported significantly more fatigue, RD=1.45% (1.20%-1.71% 95% CI), lymphadenopathy, RD=0.82% (0.62%-1.01% 95% CI), nausea, RD=0.58% (0.39%-0.78% 95% CI), headache, RD=0.40% (0.21%-0.59% 95% CI), arthralgia, RD=0.42% (0.25%-0.60% 95% CI), myalgia, RD=0.36% (0.19%-0.53% 95% CI), diarrhea, RD=0.46% (0.31%-0.62% 95% CI), fever, RD=0.30% (0.17%-0.42% 95% CI), vomiting, RD=0.30% (0.18%-0.42% 95% CI), and chills, RD=0.10% (0.02%-0.18% 95% CI) (**Figure 2**). Risk differences are also reported per vaccine brand (**Figure 2a-b**). Notably, no vaccine specific adverse events were found.

**Figure 2:**
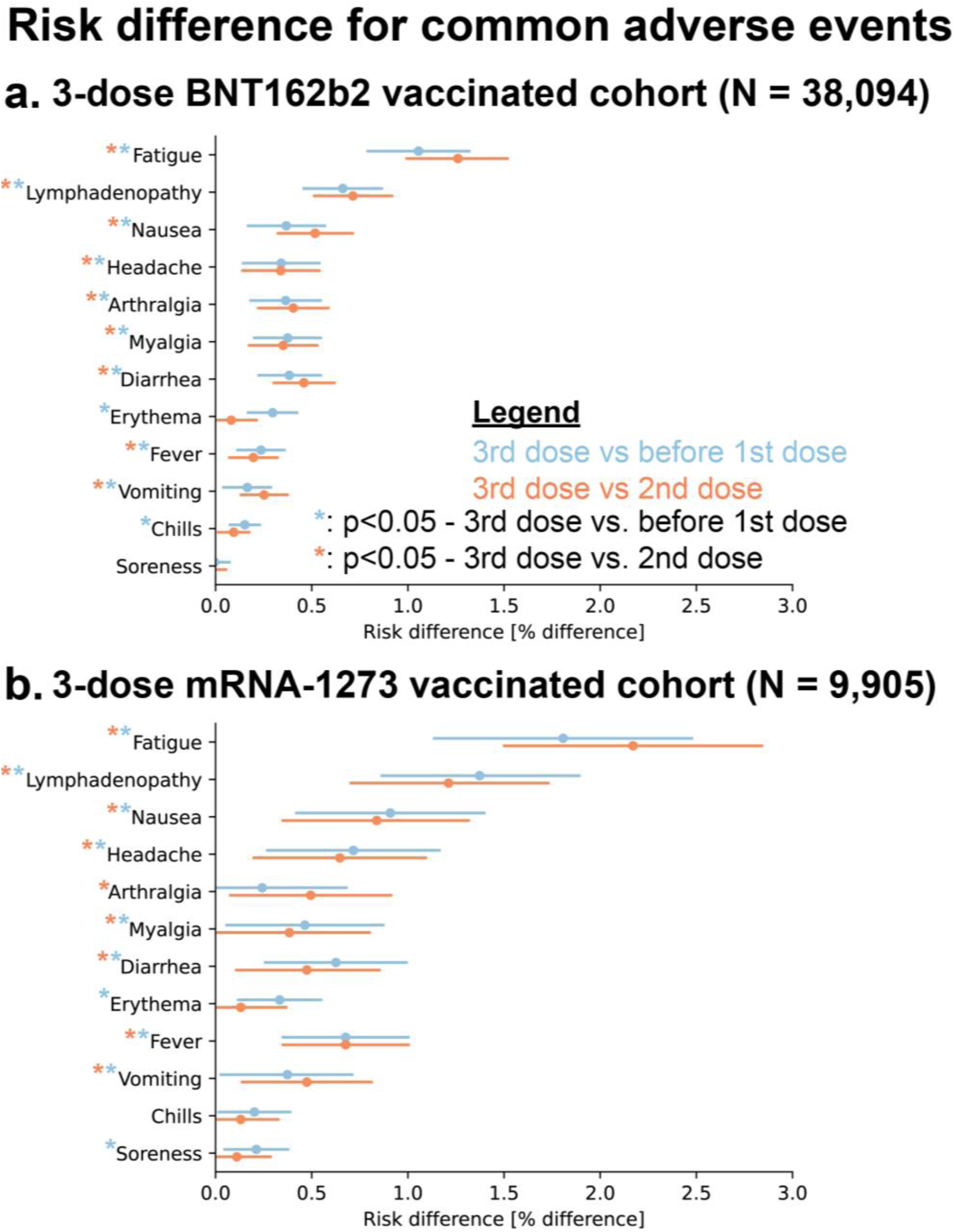
Risk difference for common adverse events after the 3^rd^ vaccine dose compared with after the 2nd dose vaccine dose (orange) and baseline risk (blue). The risk difference is shown for 3-dose BNT162b2 recipients (**a**) and for 3-dose mRNA-1273 recipients (**b**). Error bars indicate 95% confidence intervals, and asterisks indicate adverse events with significant (two-tailed p-value < 0.05) difference in prevalence after the 3^rd^ dose, compared with before the 1^st^ dose (blue asterisk) or after the 2nd dose (orange asterisk).

Reporting of severe adverse events was rare after the 3^rd^ dose and was not significantly increased compared with the frequency of those events after the 2^nd^ dose (**Figure S2**). After the 3^rd^ vaccine dose, 4 patients reported pericarditis (0.01%, 0%-0.01% 95% CI), 2 patients reported anaphylaxis (0.00%, 0%-0.01% 95% CI), and 1 patient reported myocarditis (<0.01%, 95% CI)). We find no significant increase in risk (p-value < 0.05) of these adverse events after the 3^rd^ dose, compared with after the 2^nd^ dose (**Figure S2**). Additionally, we assessed the rate of emergency department visits within 2-days of each vaccine dose and found only a slight increase after the 3^rd^ BNT162b2 dose, 0.29% of vaccine recipients (0.24%-0.35%, 95% CI) compared with after the 2^nd^ dose, 0.2% of vaccine recipients (0.15%-0.24%, 95% CI) (**Figure S3**). No significant difference in emergency department visits was found for mRNA-1273 recipients.

## Discussion

Our results suggest that a 3^rd^ dose of the same type of vaccination after either a BNT162b2 or mRNA-1273 primary series is safe. Although we observed an increase in early post-vaccination adverse events after the 3^rd^ dose compared to earlier doses, these were for symptoms of low concern (i.e., fatigue, lymphadenopathy, nausea, and diarrhea). We observed no significant increase in EHR reporting of severe adverse events after the 3^rd^ dose compared with after the 2^nd^ dose, with incidence comparable with previous literature.^16,17^ The observed increase in adverse events compared with earlier doses could be due to a stronger response elicited by the 3^rd^ dose, comparable to what was observed for the 2^nd^ dose compared with the 1^st^ dose.^1,2,33^ Further studies are needed to explore whether the 3^rd^ vaccine dose does indeed induce a stronger immune response.

This study has some limitations. To control for patient-specific covariates that may impact symptoms experienced after vaccination, we have only considered cohorts of patients that received exactly three doses of mRNA-based COVID-19 vaccines. No comparison between the adverse events reported in the BNT162b2 and mRNA-1273 cohorts should be made, as these cohorts differ in potentially confounding factors, including in the relative rate of immunosuppressed individuals (**Table S1**). The cohorts are also less likely to include individuals that had strong adverse reactions to earlier doses of mRNA-based COVID-19 vaccines, as such individuals are more likely to opt out of additional vaccination.^15^ Indeed, analysis of individuals that received exactly one dose of an mRNA-based COVID-19 vaccine showed a significantly greater RD of adverse events after dose one, compared with baseline, than what is observed in the 3-dose cohort (**Figure S4**). Additionally, a large proportion in the 3-dose cohort is immunosuppressed and of advanced age (**Table 1**), potentially reducing their immune reaction to vaccination and associated adverse events; and resulting in lower prevalence of adverse events that are more prevalent in younger individuals (i.e., myocarditis). This means that our conclusions on the safety of additional dose vaccination with BNT162b2 or mRNA-1273 apply specifically to individuals that are included in this cohort, opted to receive additional COVID-19 mRNA vaccine dose of the same type and opted to report symptoms to allow EHR capture. Therefore, these results may not be generalizable to otherwise healthy individuals. Further, we have no way of accounting for any variation in the likelihood of individuals to report outcomes; e.g. if individuals were more inclined to report certain effects after a 1^st^ vaccine dose, but are more likely to dismiss them after a 3^rd^ dose without contacting their provider, we would not be able to detect that with our data. Our eligible study population included too few individuals with mixed vaccine brands (n = 887) or with 2^nd^ doses of Ad26.COV2.S (n = 76) to reach meaningful conclusions, and these populations were therefore not included in the present report. Further study on larger more general populations might therefore find increased incidence of adverse events and will be needed to reach meaningful conclusions on the prevalence of rare adverse events.

Here, we have quantified the clinical symptoms experienced by vaccine recipients by augmented curation of EHR notes. The augmented curation process involves defining a list of symptoms and subsequently quantifying positive sentiments for these symptoms in EHR notes. As symptoms that were not explicitly included will not be quantified, the study design is not suitable for discovering adverse events that have not previously been associated with COVID-19 vaccination. Identification of positive sentiments for vaccine adverse events using augmented curation is not perfect, however previous studies have demonstrated excellent accuracy of the used augmented curation algorithms for related tasks.^28^

Extraction of adverse events from EHR notes is complementary with the clinical trials and self-reporting approaches used in previous studies. Barriers associated with self-reporting of adverse events (i.e., via a survey or device) are removed, and all adverse events for which an individual seeks medical attention will be counted. This reduction of barriers to data collection allowed us to analyze a larger cohort of 3-dose vaccine recipients than in previous studies, without necessary selection for individuals willing to self-report. However, we will only detect symptoms that individuals reported to clinicians and that were captured in EHR notes. This likely results in the low rate of common but non-severe adverse events (i.e., fatigue, local swelling, and local redness) compared with previous studies,^24,26^ as individuals may not seek medical attention for expected low-severity adverse events after vaccination.

This study provides further evidence that 3^rd^ dose vaccination with the same type of COVID-19 mRNA vaccine as used in the primary series is safe in high-risk populations. Together with previous studies of booster dose safety^13,25,26^ and effectiveness,^12,13,24^ our study supports 3^rd^ dose mRNA COVID-19 vaccination of at-risk populations.

## Data Availability

After publication, the data will be made available upon reasonable requests to the
corresponding author. A proposal with a detailed description of study objectives and the
statistical analysis plan will be needed for evaluation of the reasonability of requests.
Deidentified data will be provided after approval from the corresponding author and the Mayo
Clinic.

## Data Availability

After publication, the data will be made available upon reasonable requests to the corresponding author. A proposal with a detailed description of study objectives and the statistical analysis plan will be needed for evaluation of the reasonability of requests. Deidentified data will be provided after approval from the corresponding author and the Mayo Clinic.

## Figures

**Figure S1:**
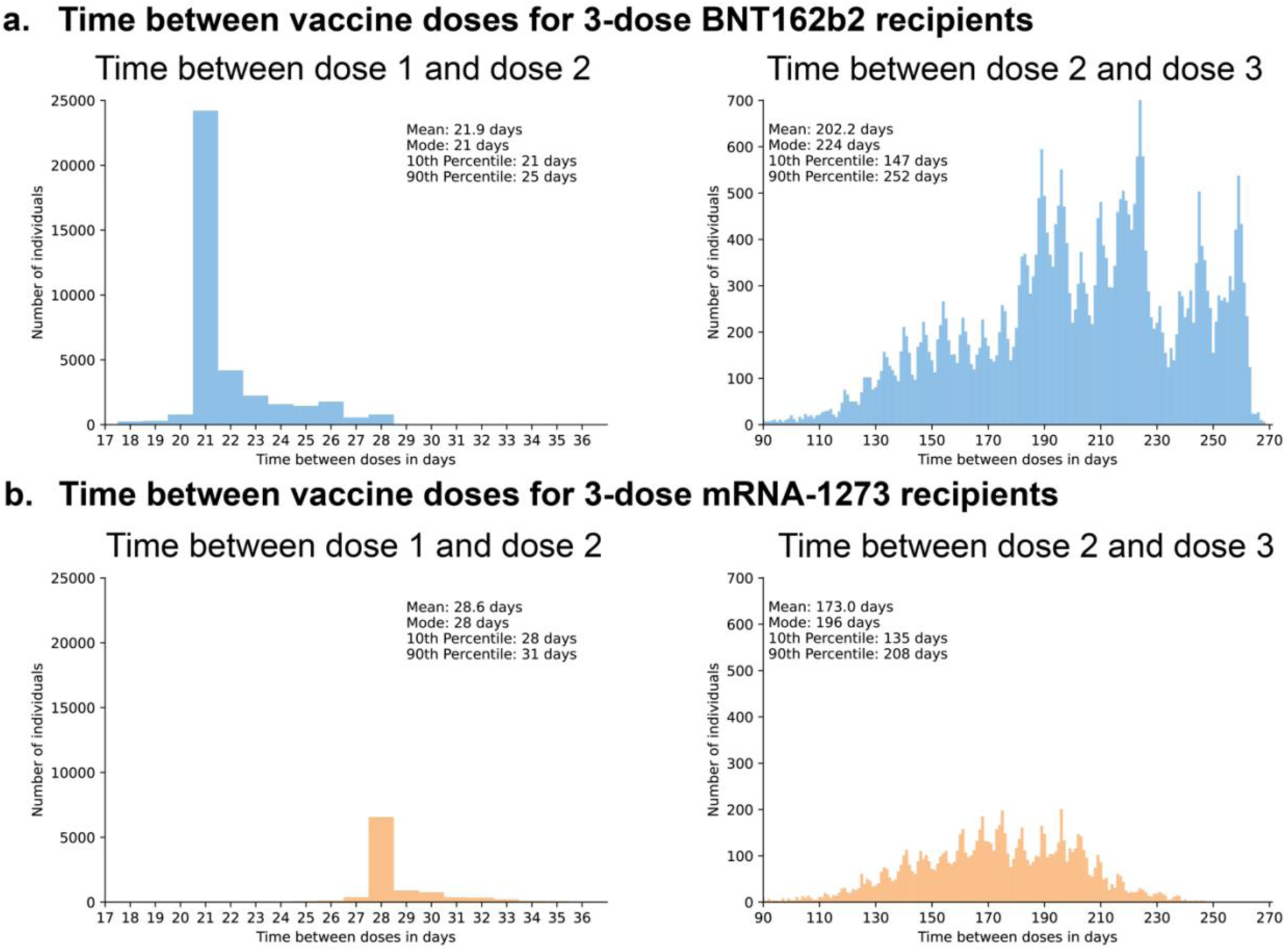
Distribution of the time between vaccine doses, for individuals that received three doses of BNT162b2 (**a)** and individuals that received three doses of mRNA-1273 (**b**).

**Figure S2:**
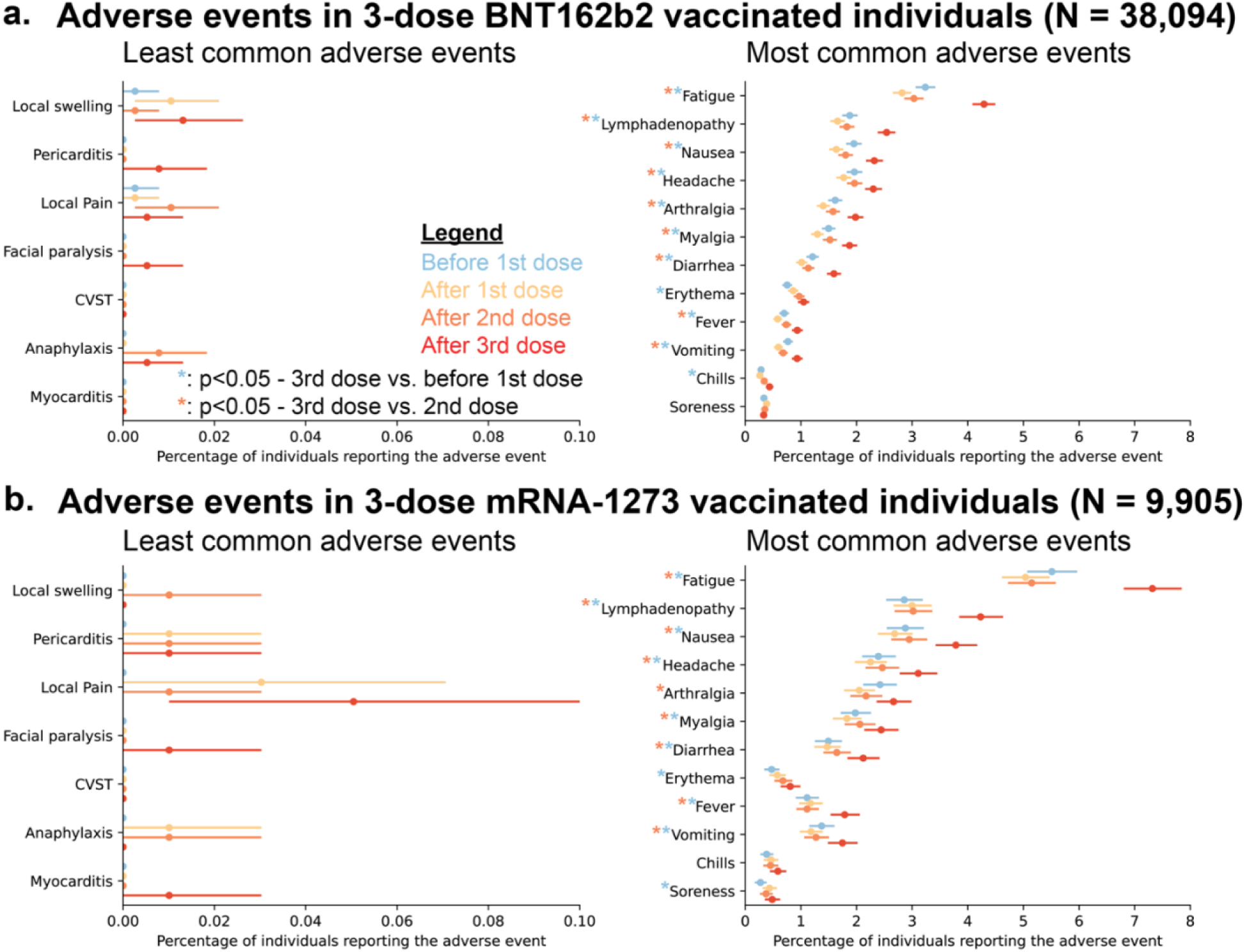
Prevalence of vaccine-associated adverse events during 14-day periods before and after vaccination in 3-dose vaccinated individuals. (**a**) Adverse events reported in individuals that received three doses of BNT162b2, before 1^st^ dose (blue), after 1^st^ dose (yellow), after 2^nd^ dose (orange), and after 3^rd^ dose (red). (**b**) Adverse events reported in individuals that received three doses of mRNA-1273, before 1^st^ dose (blue), after 1^st^ dose (yellow), after 2^nd^ dose (orange), and after 3^rd^ dose (red). Error bars indicate 95% confidence intervals, and asterisks indicate adverse events with significant (two-tailed p-value < 0.05) difference in prevalence after the 3^rd^ dose, compared with before the 1^st^ dose (blue asterisk) or after the 2^nd^ dose (orange asterisk).

**Figure S3:**
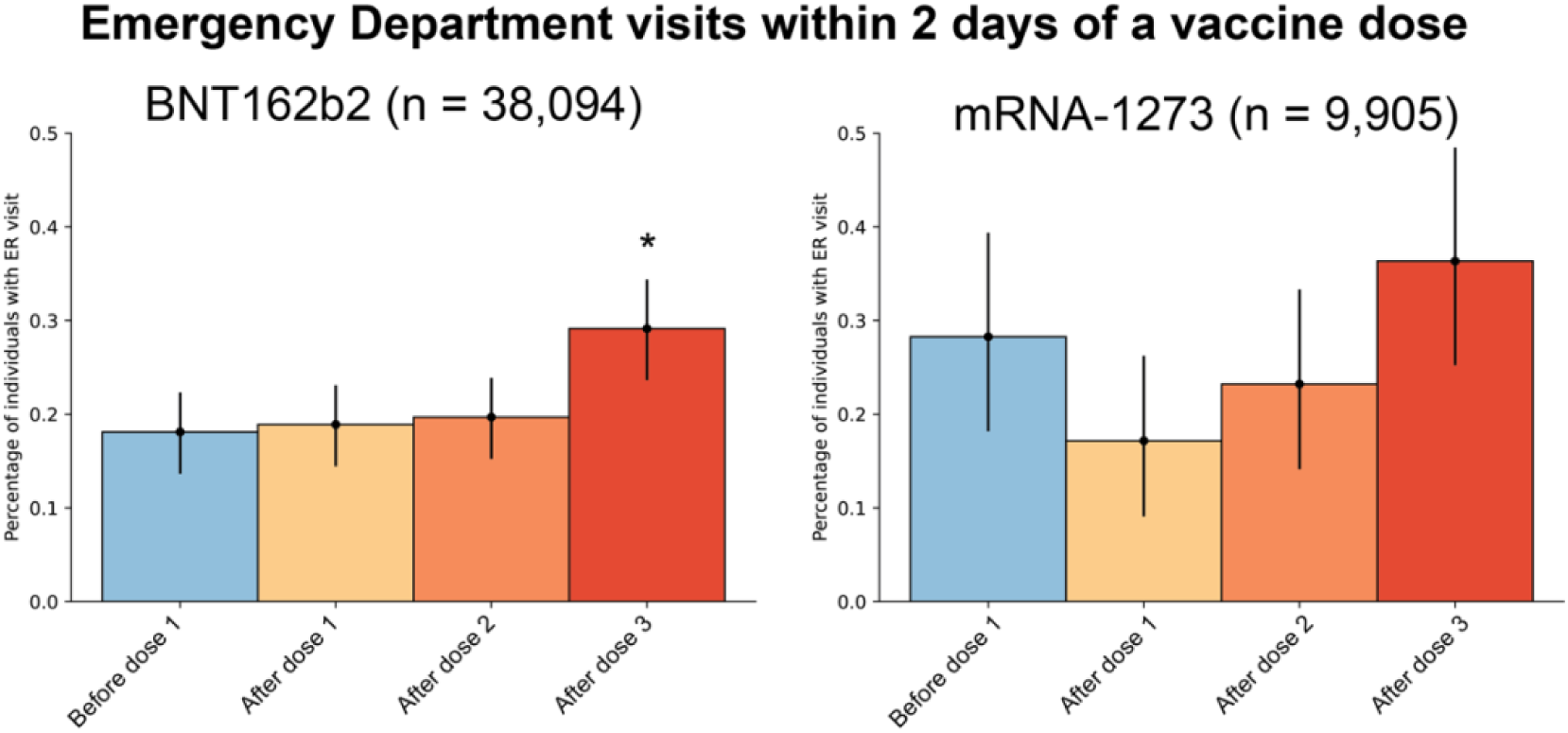
Emergency department visits within 2 days of each vaccine dose. The percentage of 3-dose mRNA vaccine recipients that visited the emergency department within 2 days after the 1^st^ dose (yellow), 2^nd^ dose (orange), or 3^rd^ dose (red), compared with a 2-day period, starting 14-days before their 1^st^ dose (blue). Error bars indicate 95% confidence intervals, and asterisks indicate a significant (two-tailed p-value < 0.05) difference in emergency department visits after vaccination compared with before.

**Figure S4:**
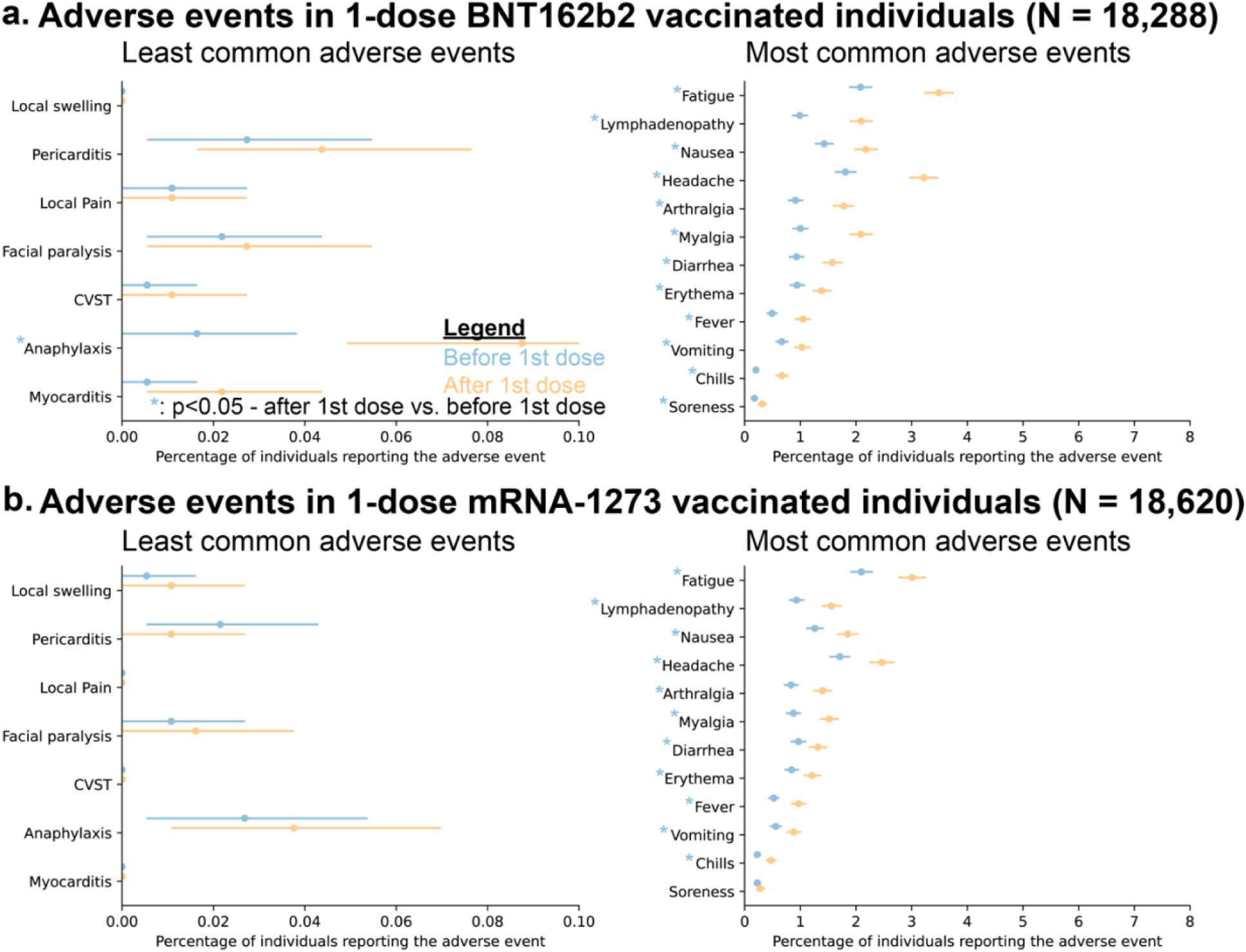
Prevalence of vaccine-associated adverse events during 14-day periods before and after vaccination in 1-dose vaccinated individuals. (**a**) Adverse events reported in individuals that received only one dose of BNT162b2, before (blue) and after (yellow) 1^st^ dose. (**b**) Adverse events reported in individuals that received only one dose of mRNA-1273, before (blue) and after (yellow) 1^st^ dose. Error bars indicate 95% confidence intervals, and asterisks indicate adverse events with significant (two-tailed p-value < 0.05) difference in prevalence after vaccination compared with before (blue asterisk).

## Tables

**Table S1:** Demographic and tracked comorbidities of the study participants.

|  | 3-dose BNT162b2 | 3-dose mRNA-1273 |
| --- | --- | --- |
| **Number of individuals, N** | 38,709 | 10,140 |
| **Sex - number (percentage)** |  |  |
| Female | 22,208 (57%) | 5,217 (51%) |
| Male | 16,495 (43%) | 4,923 (49%) |
| Unknown | 6 (<1%) | 0 |
| **Age (in years) - Mean (SD)** | 64 (17) | 65 (13) |
| **Race - number (percentage)** |  |  |
| Asian | 1,220 (3%) | 175 (2%) |
| Black/African American | 602 (2%) | 286 (3%) |
| Native American | 73 (<1%) | 43 (<1%) |
| Hawaiian/Pacific Islander | 24 (<1%) | 5 (<1%) |
| White/Caucasian | 35,777 (92%) | 9,396 (93%) |
| Other | 596 (2%) | 128 (1%) |
| Unknown | 417 (1%) | 107 (1%) |
| **Ethnicity - number (percentage)** |  |  |
| Hispanic or Latino | 893 (2%) | 303 (3%) |
| Not Hispanic or Latino | 36,848 (95%) | 9,603 (95%) |
| Unknown | 968 (3%) | 234 (2%) |
| **Elixhauser index - Mean (SD)** | 7.6 (10.9) | 12.1 (12.7) |
| **Comorbidities - number (percentage)** |  |  |
| Immunosuppressed (within 1 year) | 4,860 (13%) | 2,897 (29%) |
| Cancer | 8,402 (22%) | 3,435 (34%) |
| HIV/AIDS | 159 (<1%) | 90 (<1%) |

**Table S2:** Number of reports for tracked adverse events in each analysis time-window with median time, in days, between report and date of vaccination, for the 3-dose BNT162b2 cohort.

| **Adverse event** | **#reports**<br>**after 1<sup>st</sup>**<br>**dose of**<br>**BNT162b2** | **Median**<br>**days**<br>**between 1<sup>st</sup>**<br>**dose and**<br>**report** | **#reports**<br>**after 2<sup>nd</sup>**<br>**dose of**<br>**BNT162b2** | **Median**<br>**days**<br>**between**<br>**2<sup>nd</sup> dose**<br>**and report** | **#reports**<br>**after 3<sup>rd</sup>**<br>**dose of**<br>**BNT162b2** | **Median**<br>**days**<br>**between 3<sup>rd</sup>**<br>**dose and**<br>**report** | **#reports**<br>**before 1<sup>st</sup>**<br>**dose of**<br>**BNT162b2** |
| --- | --- | --- | --- | --- | --- | --- | --- |
| Anaphylaxis | 0 | not reported | 3 | 9 | 2 | 3.5 | 0 |
| Arthralgia | 534 | 6 | 601 | 7 | 755 | 4 | 616 |
| CVST | 0 | not reported | 0 | not reported | 0 | not reported | 0 |
| Chills | 100 | 8 | 130 | 6 | 166 | 6 | 108 |
| Diarrhea | 386 | 7 | 432 | 6 | 607 | 5 | 461 |
| Erythema | 328 | 7 | 369 | 6 | 400 | 6 | 287 |
| Facial paralysis | 0 | not reported | 0 | not reported | 2 | 6.5 | 0 |
| Fatigue | 1,074 | 7 | 1,155 | 7 | 1635 | 4 | 1233 |
| Fever | 222 | 7.5 | 281 | 7 | 356 | 6 | 266 |
| Headache | 674 | 7 | 748 | 6 | 877 | 5 | 747 |
| Local pain | 1 | 8 | 4 | 4 | 2 | 3.5 | 1 |
| Local swelling | 4 | 4 | 1 | 2 | 5 | 4 | 1 |
| Lymphadenopathy | 632 | 7 | 696 | 7 | 968 | 3 | 716 |
| Myalgia | 494 | 6 | 580 | 7 | 714 | 4 | 571 |
| Myocarditis | 0 | not reported | 0 | not reported | 0 | not reported | 0 |
| Nausea | 623 | 7 | 687 | 7 | 884 | 6 | 744 |
| Pericarditis | 0 | 7 | 0 |  | 3 | 4 | 0 |
| Soreness | 146 | 7 | 134 | 7 | 126 | 5 | 128 |
| Vomiting | 227 | 7 | 259 | 7 | 355 | 5 | 292 |

**Table S3:**
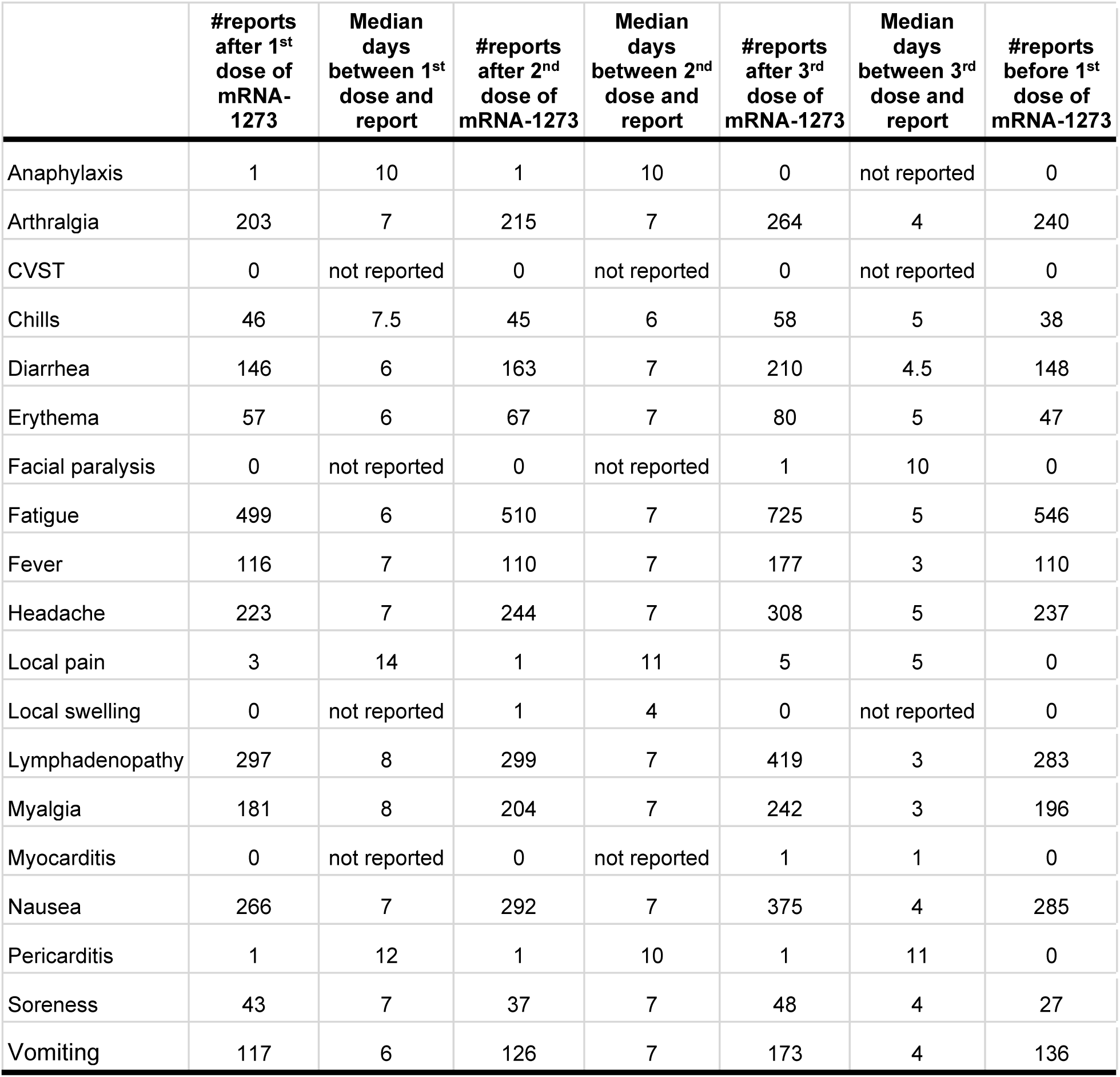
Number of reports for tracked adverse events in each analysis time-window with median time, in days, between report and date of vaccination, for the 3-dose mRNA-1273 cohort.

|  | #reports<br>after 1<sup>st</sup><br>dose of<br>mRNA-1273 | Median<br>days<br>between 1<sup>st</sup><br>dose and<br>report | #reports<br>after 2<sup>nd</sup><br>dose of<br>mRNA-1273 | Median<br>days<br>between 2<sup>nd</sup><br>dose and<br>report | #reports<br>after 3<sup>rd</sup><br>dose of<br>mRNA-1273 | Median<br>days<br>between 3<sup>rd</sup><br>dose and<br>report | #reports<br>before 1<sup>st</sup><br>dose of<br>mRNA-1273 |
| --- | --- | --- | --- | --- | --- | --- | --- |
| Anaphylaxis | 1 | 10 | 1 | 10 | 0 | not reported | 0 |
| Arthralgia | 203 | 7 | 215 | 7 | 264 | 4 | 240 |
| CVST | 0 | not reported | 0 | not reported | 0 | not reported | 0 |
| Chills | 46 | 7.5 | 45 | 6 | 58 | 5 | 38 |
| Diarrhea | 146 | 6 | 163 | 7 | 210 | 4.5 | 148 |
| Erythema | 57 | 6 | 67 | 7 | 80 | 5 | 47 |
| Facial paralysis | 0 | not reported | 0 | not reported | 1 | 10 | 0 |
| Fatigue | 499 | 6 | 510 | 7 | 725 | 5 | 546 |
| Fever | 116 | 7 | 110 | 7 | 177 | 3 | 110 |
| Headache | 223 | 7 | 244 | 7 | 308 | 5 | 237 |
| Local pain | 3 | 14 | 1 | 11 | 5 | 5 | 0 |
| Local swelling | 0 | not reported | 1 | 4 | 0 | not reported | 0 |
| Lymphadenopathy | 297 | 8 | 299 | 7 | 419 | 3 | 283 |
| Myalgia | 181 | 8 | 204 | 7 | 242 | 3 | 196 |
| Myocarditis | 0 | not reported | 0 | not reported | 1 | 1 | 0 |
| Nausea | 266 | 7 | 292 | 7 | 375 | 4 | 285 |
| Pericarditis | 1 | 12 | 1 | 10 | 1 | 11 | 0 |
| Soreness | 43 | 7 | 37 | 7 | 48 | 4 | 27 |
| Vomiting | 117 | 6 | 126 | 7 | 173 | 4 | 136 |

